# In-hospital Outcomes of Septal Myectomy Versus Alcohol Septal Ablation for Hypertrophic Cardiomyopathy with Outflow Tract Obstruction - An Update and Insights from The National Inpatient Sample from 2011-2019

**DOI:** 10.1101/2023.06.07.23291116

**Authors:** Karla Inestroza, Ivan Mijares-Rojas, Carlos Matute-Martínez, Ian Ergui, Michael Albosta, Carlos Vergara-Sanchez, Michael Dangl, Rafael Jaciel Hernandez, Bertrand Ebner, Louis T Vincent, Jelani Grant, Jennifer Maning, Carlos Alfonso, Rosario Colombo

## Abstract

**Background:** Septal Myectomy (SM) and Alcohol Septal Ablation (ASA) improve symptoms in patients with Hypertrophic Cardiomyopathy with outflow tract obstruction (oHCM). However, outcomes data in this population is predominantly from specialized centers.

**Methods:** The National Inpatient Database was queried from 2011- 2019 for relevant ICD-9 and −10 diagnostic and procedural codes. We compared baseline characteristics and in-hospital outcomes of patients with oHCM who underwent SM vs. ASA. A p-value <0.001 was considered statistically significant.

**Results:** We identified 17,245 patients with oHCM who underwent septal reduction therapies, of whom 62.5% underwent SM, and 37.5% underwent ASA. Patients who underwent SM had higher all-cause mortality (OR:2.2 [1.7-2.9]), post-procedure ischemic stroke (OR: 2.4 [1.8-3.2]), acute kidney injury (OR: 1.9 [1.7-2.2]), vascular complications (OR: 4 [2.8-5.7]), ventricular septal defect (OR: 4.6 [3.5-6.1]), cardiogenic shock (OR: 2 [1.5-2.6]), sepsis (OR: 5.2 [3.3-8.1]), and left bundle branch block (OR: 3.2 [2.8-3.7]), compared to ASA. Patients who underwent ASA had higher post-procedure complete heart block (OR: 1.2 [1.1-1.4]), 2nd-degree AV Block (OR: 2 [1.4-3]), right bundle branch block (OR: 6.4 [5.3-7.8]), ventricular tachycardia (OR:2 [1.8-2.3]), supraventricular tachycardia (OR: 1.4 [1.2-1.7]), and more commonly required pacemaker (OR: 1.4 [1.2-1.6]) or implantable cardioverter-defibrillator insertion (OR: 1.3 [1.1-1.5]) (p<0.001 for all) compared to SM.

**Conclusions:** This nationwide analysis evidenced that patients undergoing SM had higher in-hospital mortality and periprocedural complications than ASA; however, those undergoing ASA had more post-procedure conduction abnormalities and pacemaker or ICD implantation. The implications of these findings warrant further investigation regarding patient selection strategies for these therapies.

## Introduction

Hypertrophic cardiomyopathy (HCM) is the most common inheritable heart disease, with an approximate prevalence of 1 case in 500 persons in the general population.^1^ In the past 50 years, its perception in the cardiovascular field has changed from a rare, untreatable condition with an obscure prognosis to a disease with multimodal therapeutic approaches and a heterogeneous clinical presentation. HCM is inherited in an autosomal dominant pattern with mutations involving genes encoding proteins of the contractile myofilament apparatus.^2^ Histopathologically, it is identified by cardiomyocyte hypertrophy, disarray, and increased fibrosis.^3^

Clinical presentation includes sudden cardiac death (SCD), sometimes as the initial manifestation, chronic Atrial Fibrillation (A. Fib), and symptomatic Heart Failure (HF), with many patients having a degree of left ventricular outflow tract obstruction (LVOTO).^1^

The current therapeutic approach emphasizes SCD prevention with an implantable cardiac defibrillator (ICD).^1, 4, 5^ Progressive heart failure symptomatology is treated with lifestyle modifications to avoid hemodynamic LVOTO and atrioventricular nodal blocking agents (beta-adrenergic and calcium channel blockers) or disopyramide in exceptional cases.^4, 6^

Septal reduction therapy (SRT) is the next available option for a particular subset of patients that are refractory to drug therapy with a left ventricular outflow tract (LVOT) gradient ≥50 mmHg (at rest or provoked) and impaired quality of life.^1, 7^ The surgical approach with Septal Myectomy (SM) reduces the degree of LVOTO and improves cardiac function and prognosis.^4, 5, 7, 8^ However, the less invasive interventional approach with Alcohol Septal Ablation (ASA) has also proved to be a safe alternative for patients with high surgical risk, and both procedures have a Class 1 recommendation as per the 2020 AHA/ACC Guideline for the Diagnosis and Treatment of Patients With Hypertrophic Cardiomyopathy.^4, 5^

The present study compared available data from the National Inpatient Sample (NIS) to illuminate safety endpoints and outcomes between SRT modalities.

## Methods

### Data Source

The NIS offers the largest database of hospitalizations, including data on approximately 7-8 million discharges per year and representing a 20% random and stratified sample of hospital discharges in the United States. Annual data quality assessments of the NIS are performed, guaranteeing the database’s internal validity. We used the International Classification of Diseases Ninth Revision, Clinical Modification (ICD-9-CM) and International Classification of Diseases Tenth Revision, Clinical Modification (ICD-10- CM) diagnostic and therapeutic procedure codes to identify the study population. Institutional review board approval was not needed as all patient information is de-identified within the NIS.

### Data Disclosure

All the data under NIS are publicly available. Detailed ICD diagnostic and procedural codes used for statistical analyses are presented under the Data Supplement, which can be used to replicate our results.

### Study Population

We performed a retrospective analysis using the NIS database from January 1, 2011, to December 31, 2019. We identified all patients aged≥18 with a diagnosis of HCM with outflow tract obstruction (oHCM) using the International ICD-9-CM Revision-Diagnostic Coding System code 425.11 or ICD-10-CM Revision-Diagnostic Coding System code I42.1 in any of the first ten diagnostic fields, identifying a total of 51,877 cases. Then, we proceeded to identify patients who underwent SM using the International ICD-9-CM Revision-Procedure Coding System code 37.33 or ICD-10-CM Revision-Procedure Coding System code 02BM, 02BM0, 02BM0Z, and 02BM0ZZ in any of the first five diagnostic fields, and patients who underwent ASA using international ICD-9-CM Revision-Procedure Coding System code 37.34 or ICD-10-CM Revision-Procedure Coding System code 025M3, 025M3Z, 025M3ZZ, 02583, 02583Z, or 02583ZZ in any of the first five diagnostic fields. Our final study cohort comprised 17,245 patients with oHCM who underwent either SM (10,781 patients) or ASA (6,464 patients). International Classification of Diseases coding for comorbidities and post-SRT (SM or ASA) complications are listed in Table 1 in the Data Supplement.

**Table 1.** Baseline characteristics of patients with Hypertrophic Cardiomyopathy with Outflow Tract Obstruction who underwent Septal Myectomy vs. Alcohol Septal Ablation.

| **Variable** | **Septal Myectomy**<br>**(N=10,781)** | **Alcohol Septal**<br>**Ablation (N=6,464)** | **P-value\*** |
| --- | --- | --- | --- |
| **Baseline characteristics** |  |  |  |
| Age in years | 58.9 ± 14.1 | 63.3 ± 13.6 | <0.001 |
| Female Gender | 4,805 (44.6%) | 2,960 (45.8%) | 0.118 |
| White Race | 9,039 (83.8%) | 5,161 (79.8%) | <0.001 |
| Hypertension | 7,450 (72%) | 4,475 (72.2%) | 0.960 |
| Diabetes Mellitus | 1,895 (17.7%) | 1,213 (18.8%) | 0.055 |
| Atrial Fibrillation | 4,643 (43%) | 3,058 (47.3) | <0.001 |
| Atrial Flutter | 688 (6.4%) | 1,325 (18.9%) | <0.001 |
| Tobacco use | 4,025 (37.3%) | 1,992 (30.8%) | <0.001 |
| Alcohol use | 243 (2.3%) | 84 (1.3%) | <0.001 |
| Substance use | 165 (1.5%) | 45 (0.7%) | <0.001 |
| Hyperlipidemia | 5,784 (57.2%) | 3,477 (57.7%) | 0.508 |
| Obesity | 3,148 (29.2%) | 1,229 (19%) | <0.001 |
| Anemia | 299 (3.4%) | 194 (3.8%) | <0.001 |
| Obstructive Sleep Apnea | 2,200 (20.4%) | 1,349 (20.9%) | 0.468 |
| Presence of pacemaker | 359 (3.3%) | 521 (8.1%) | <0.001 |
| Presence of an Implantable<br>Cardioverter Defibrillator | 1,343 (12.5%) | 1,353 (20%) | <0.001 |
| Thrombocytopenia | 2,735 (25.4%) | 238 (3.7%) | <0.001 |
| Heart Failure | 4,568 (42.5%) | 2,557 (39.6%) | <0.001 |
| All prior strokes | 526 (3.95%) | 503 (7.8%) | <0.001 |
| Coronary Artery Disease | 3,832 (35.9%) | 2,233 (34.7%) | <0.001 |
| Peripheral Artery Disease | 418 (4%) | 165 (2.6%) | <0.001 |
| Chronic Kidney Disease | 1,126 (10.5%) | 805 (12.5%) | <0.001 |
| Chronic Obstructive<br>Pulmonary Disease | 1,159 (10.9%) | 842 (13.2%) | <0.001 |
| History of Myocardial<br>Infarction | 476 (4.4%) | 416 (6.4%) | <0.001 |
| Liver Disease and Cirrhosis | 264 (2.5%) | 200 (3.1%) | 0.014 |
| History of Percutaneous Coronary Intervention | 654 (6%) | 632 (9.8%) | <0.001 |
| History of Coronary Artery Bypass Graft | 129 (1.2%) | 140 (2.2%) | <0.001 |
| Pulmonary Hypertension | 1,522 (14.1%) | 644 (9.9%) | <0.001 |

\* A p-value <0.001 was considered statistically significant.

**Table 2.**
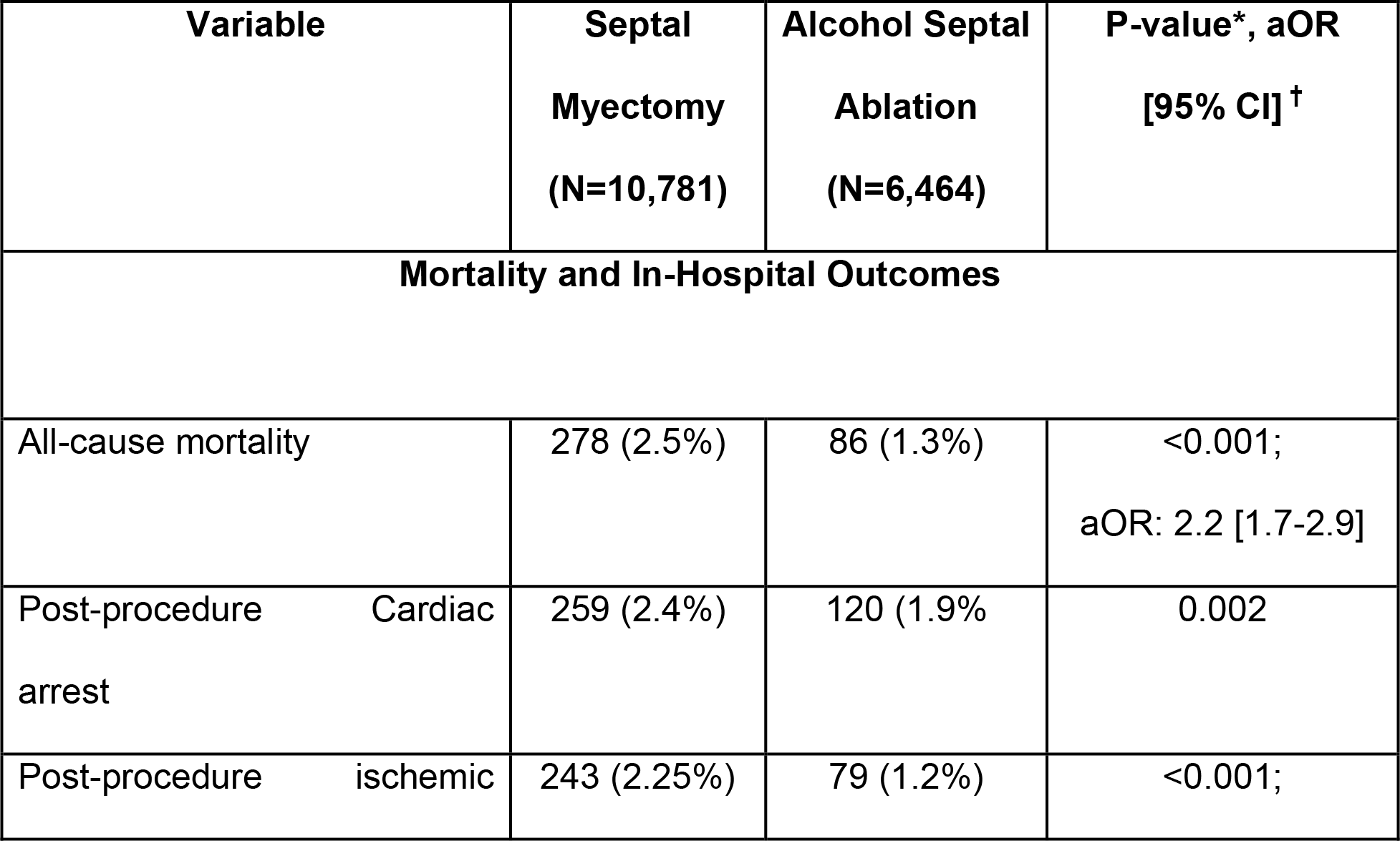
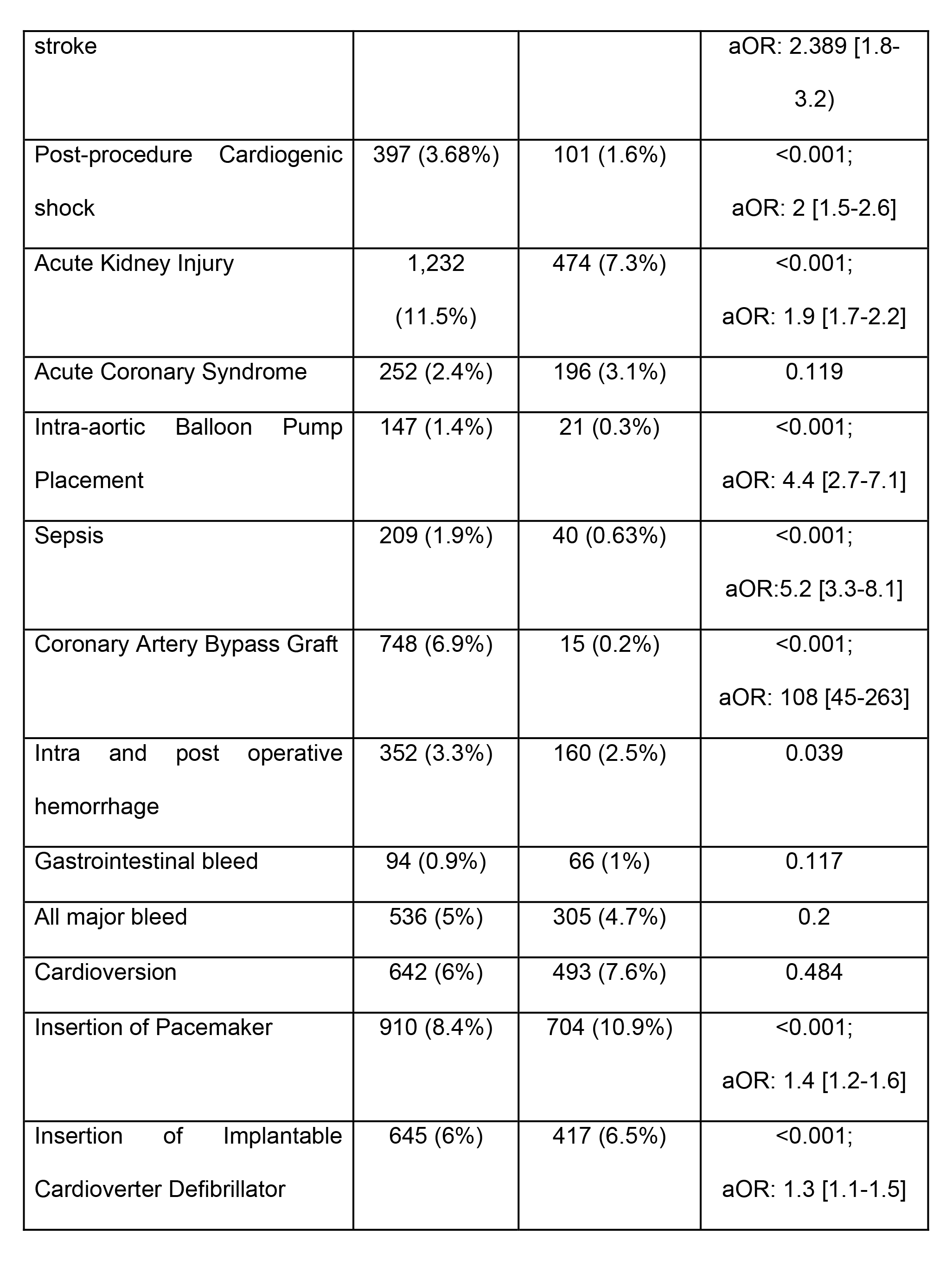
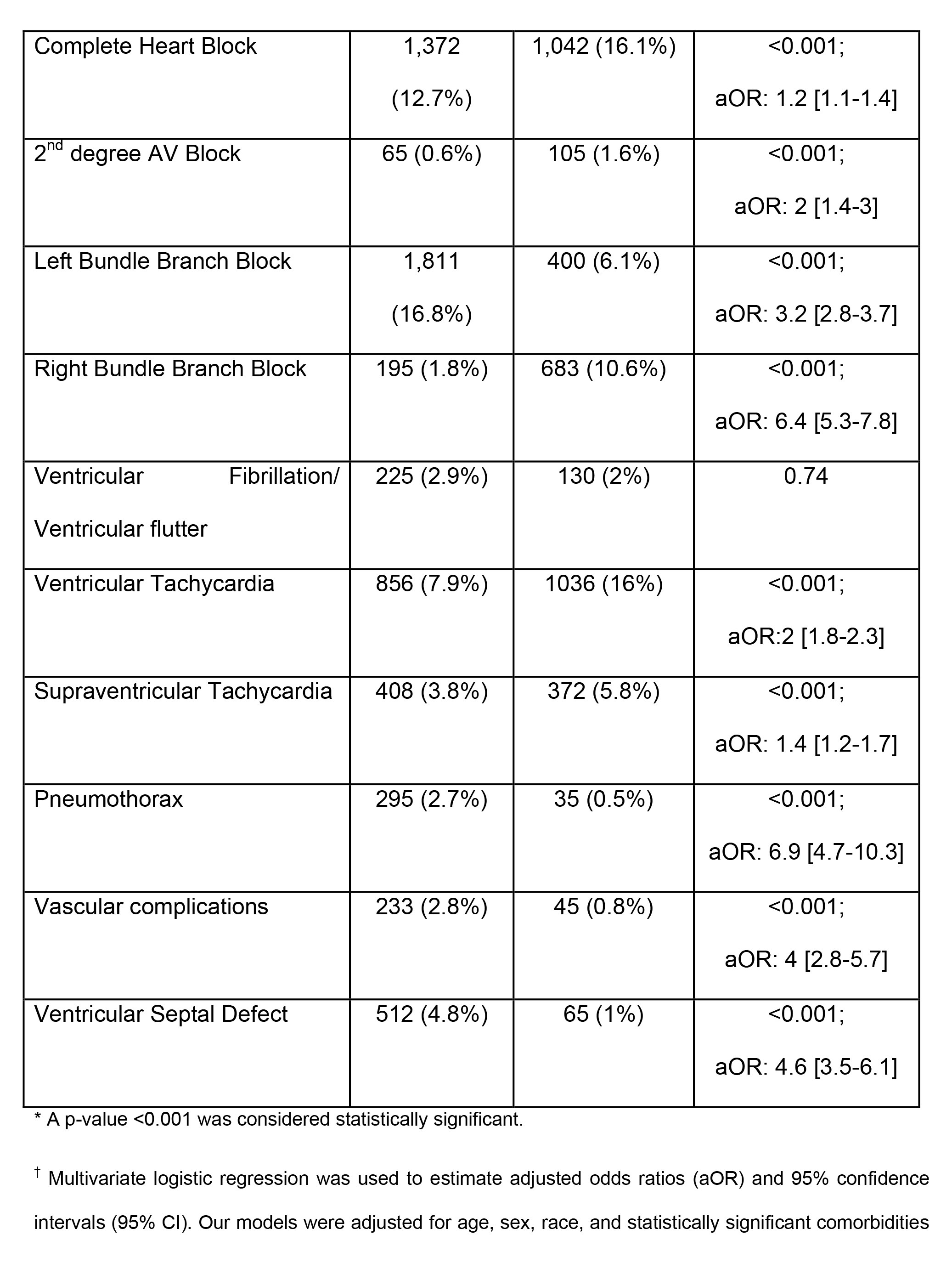
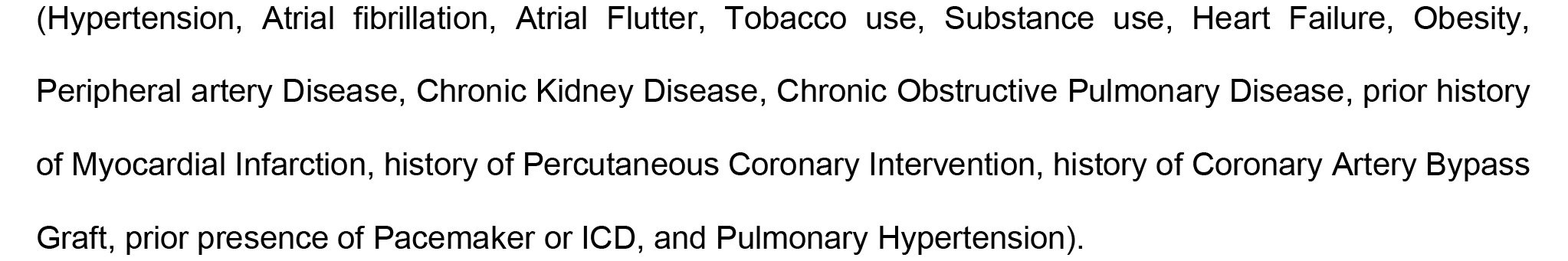
Mortality and In-hospital outcomes in patients with Hypertrophic Cardiomyopathy with Outflow Tract Obstruction who underwent Septal Myectomy vs. Alcohol Septal Ablation.

### Outcome Measures

Our primary outcome was post-SRT (SM or ASA) all-cause In-hospital mortality.

Secondary outcomes of interest were post-procedure complications with cardiac arrest, ischemic stroke, cardiogenic shock, acute kidney injury (AKI), sepsis, major vascular complications (aortic dissection, aortic aneurysm, postoperative deep vein thrombosis [DVT] or pulmonary embolism [PE]), bleeding, need for insertion of a pacemaker or ICD, cardioversion, conduction abnormalities (complete heart block, 2nd-degree atrioventricular block, and left or right bundle branch block), and arrhythmias (Ventricular fibrillation/flutter, Ventricular tachycardia, Supraventricular Tachycardia [SVT]) for indexed hospitalizations during which patients underwent SM or ASA.

### Statistical Analysis

As the Agency for Healthcare Research and Quality recommended, weighted data were used for all statistical analyses. Baseline characteristics and post-procedure outcomes were compared using the Pearson Chi-Squared (χ²) tests for categorical variables and Independent Sample T-testing for continuous variables. A p-value <0.001 was considered statistically significant.

Multivariate logistic regression was used to estimate adjusted odds ratios (aOR) and 95% confidence intervals (95% CI) to determine the association between oHCM and post-SRT (SM vs. ASA) mortality and In-hospital outcomes. All multivariate logistic regression models were created using generalized estimating equations. Our models were adjusted for age, sex, race, and statistically significant comorbidities (HTN, A. Fib, Atrial Flutter, Tobacco use, Substance use, Heart HF, Obesity, PAD, CKD, Chronic Obstructive Pulmonary Disease (COPD), prior history of Myocardial Infarction [MI], history of Percutaneous Coronary Intervention [PCI], history of Coronary Artery Bypass Graft [CABG], prior presence of Pacemaker or ICD, and Pulmonary Hypertension [PH]) between groups. As recommended by Healthcare Cost and Utilization Project, missing data for age, gender, and race were handled using multiple imputations. All statistical analyses were performed using SPSS (IBM SPSS Statistics for Mac, Version 23.0. Armonk, NY: IBM Corp.).

## Results

### Baseline Patient Characteristics

We identified 17,245 patients with oHCM who underwent septal reduction procedures during the years 2011-2019, of whom 10,781 (62.5%) underwent SM and 6,464 (37.5%) underwent ASA. The average age of the cohort was 60.5 years ± 14.1 years, with a median age of 62 years, ranging from 18 to 92 years. The female sex accounted for 45% (7,764) of the total population. Regarding race, white individuals accounted for the majority (82.3%) of the total population.

All comparisons henceforth are reported as SM versus (vs.) ASA for consistency. Individuals who underwent SM were younger 58.9 ± 14.1 years vs. 63.3 ± 13.6 years (p<0.001), had a lower prevalence of A. Fib (43% vs. 47.3%, p<0.001), Atrial Flutter (6.4% vs. 18.9%, p<0.001), and of prior presence of a pacemaker (3.3% vs. 8.1%, p <0.001) or ICD (12.5% vs. 20%, p<0.001), lower history of prior stroke (3.95% vs. 7.8%, p<0.001), CKD (10.5% vs. 12.5%, p<0.001), COPD (10.9% vs. 13.2%, p<0.001), history of prior MI (4.4% vs. 6.4%, p<0.001), history of previous PCI (6% vs. 9.8%, p<0.001) and prior CABG (1.2% vs. 2.2%, p<0.001) compared to those who underwent ASA.

Conversely, those who underwent SM had higher rates of tobacco use (37.3% vs. 30.8%, p<0.001), alcohol use (2.3% vs. 1.3%, p<0.001), and substance use (1.5% vs. 0.7%, p<0.001), and higher prevalence of obesity (29.2% vs. 19%, p<0.001), more HF (42.5% vs. 39.6%, p<0.001), more CAD (35.9% vs. 34.7%, p<0.001), more PAD (4% vs. 2.6%, p<0.001), and more PH (14.1% vs. 9.9%, p<0.001) compared to those who underwent ASA.

There was no statistically significant difference in sex between groups, with the female gender compromising 44.6% vs. 45.8% (p=0.118) of the population undergoing SM vs. ASA respectively. The incidence of HTN (72% vs. 72.2%, p=0.960), DM (17.7% vs. 18.8%, p=0.055), Hyperlipidemia (HLD, 57.2% vs. 57.7%, p=0.508), Obstructive Sleep Apnea (OSA, 20.4% vs. 20.9, p= 0.468), and liver disease and cirrhosis (2.5% vs. 3.1%, p=0.014) was not statistically different between the compared groups.

### In-hospital outcomes and procedural complications

In regards to inpatient post-procedure complications, patients who underwent SM had higher rates of post-procedure ischemic stroke (2.25% vs. 1.2%, p<0.001; aOR:2.4 [95% CI:1.8-3.2]), AKI (11.5% vs. 7.3%, p<0.001; aOR:1.9 [95% CI:1.7-2.2]), VSD (4.8% vs. 1%, p<0.001; aOR:4.6 [95% CI:3.5-6.1]), cardiogenic shock (3.68% vs. 1.6%, p<0.001; aOR:2 [95% CI:1.5-2.6]), sepsis (1.9% vs. 0.63%, p<0.001; aOR:5.2 [95% CI:3.3-8.1]), vascular complications (2.8% vs. 0.8%, p<0.001; aOR:4 [95% CI:2.8-5.7]), pneumothorax (2.7% vs. 0.5%, p<0.001; aOR:6.9 [95% CI:4.7-10.3]), Left Bundle Branch Block (LBBB, 16.8% vs. 10.6%, p<0.001; aOR:3.2 [95% CI:2.8-3.7]), and more frequently required Intra-aortic balloon pump placement (IABP) placement (1.4% vs. 0.3%, p<0.001; aOR:4.4 [95% CI:2.7-7.1]) compared to ASA.

Compared to patients who underwent SM, those who underwent ASA had higher post-procedure complete heart block (12.7% vs. 16.1%, p<0.001; aOR:1.2 [95% CI:1.1-1.4]), 2nd-degree AV Block (0.6% vs. 1.6%, p<0.001; aOR:2 [95% CI:1.4-3]), Right Bundle Branch Block (RBBB, 1.8% vs. 10.6%, p<0.001; aOR:6.4 [95% CI:5.3-7.8]), ventricular tachycardia (7.9% vs. 16%, p<0.001; aOR:2 [95% CI:1.8-2.3]), SVT (3.8% vs. 5.8%, p<0.001; aOR:1.4 [95% CI:1.2-1.7]), and more commonly required pacemaker (8.4% vs. 10.9%, p<0.001; aOR:1.4 [95% CI:1.2-1.6]) or ICD insertion (6% vs. 6.5%, p<0.001; aOR:1.3 [95% CI:1.1-1.5]).

There was no difference in rates of cardiac arrest (2.4% vs. 1.9%, p=0.002), ACS (2.4% vs. 3.1%, p=0.119), Ventricular fibrillation/Ventricular Flutter (2.9% vs. 2%, p<0.74) intra-operative and post-operative bleeding (3.3% vs. 2.5%, p=0.039), Gastrointestinal bleeding (0.9% vs. 1%, p=0.117), or requiring cardioversion (6% vs. 7.6%, p=0.484) between the compared groups.

### Association between post-procedure SM vs. ASA inpatient mortality

Combining data from the studied years (2011-2019), patients undergoing SM had higher inpatient all-cause mortality (2.5% vs. 1.3%, p<0.001) that remained significant beyond adjustment for demographic factors and comorbidities (aOR:2.2 [95% CI:1.7- 2.9]).

## Discussion

This nationwide retrospective analysis studying the outcomes after SM and ASA in patients with oHCM from 2011 to 2019 evidenced several essential findings that intend to better prepare the physician and the patient on the unique post-procedure expectations and the most common complications of each septal reduction intervention, thus facilitating and providing a basis for shared decision-making at the time of procedure and patient selection.

First, we found that the all-cause mortality after SM (2.5%) was higher than prior reports from single center-high volume experiences, where low postoperative mortality (<1%)^5,9,10,11,12^ is described, and similar to the volume-unadjusted report from the Society of Thoracic Surgeons Adult Cardiac Surgery Database^13^. We report a post-ASA mortality rate of 1.3%, which is similar to prior reports from the Mayo Clinic (1.4%)^14^ and higher than studies from the Euro-Alcohol Septal Ablation Registry (0.6%)^15^.

Our data supports findings suggesting that invasive SRTs are associated with increased mortality when performed in centers with limited experience and low procedural volume.^5,13,15,16^

We also describe a higher incidence of post-procedure complications after SM, including ischemic stroke, AKI, VSD, cardiogenic shock, requiring IABP support, sepsis, vascular complications, LBBB, and pneumothorax compared to ASA.

One of the common complications after ASA is new onset conduction abnormalities^17, 18^, which we also describe and can be attributed to the nature of the procedure itself. Ventricular tachycardia was the most common arrhythmia reported (16%). The second most common arrhythmia was the development of a RBBB- attributed to the superior location of the fibers in the ventricular septum supplied by the first septal perforator-although we found the incidence of the latter to be lower (10.6%) than reported in prior studies (58-68%)^19, 20^. Since RBBB commonly occurs in cases of ASA^17^, the risk of complete Atrio Ventricular Block (AVB) is highest in patients with preexisting LBBB.

As previously described^17, 18^, we found a higher post-ASA need for pacemaker or ICD implantation (combined 17.4%).

Prior concerns regarding ventricular fibrillation/flutter related to post-ASA septal scar are not reflected in more recent reports^5, 21, 22^, results which we also describe, reporting no difference (2.9% vs. 2%, p=0.74) when comparing both procedures.

There are several limitations present in our study: 1. This retrospective study from a large inpatient database can lead to underrepresentation or overrepresentation of the volume of procedures or co-morbid diagnosis. 2. The NIS database provides only in-hospital outcomes; thus, our findings do not reflect the long-term hemodynamic or clinical outcomes or the need for additional hospitalizations or procedures. 3. We cannot assess for LVOT gradient or improvement in symptoms after the procedure. 4. Our analysis included data from multiple hospitals nationwide that is not stratified according to procedure volume as done in reports from specialized centers.

We recognize the above limitations and that many HCM recommendations are based on observational data or expert opinion. Comparing both procedures requires larger randomized trials, which might not be feasible considering the low event rates and slow disease progression in HCM. Thus, reports from larger datasets like the NIS can be useful in providing insight on the pros and cons of the available SRT procedures.

## Conclusion

This nationwide analysis reports that patients undergoing SM had higher in-hospital mortality and periprocedural complications than those undergoing ASA. Patients who underwent ASA had more post-procedure conduction abnormalities and pacemaker or ICD implantation requirements. Comparing both procedures with larger randomized trials might not be feasible, and reports from larger datasets can provide insight into post-procedure expectations, expected complications of septal reduction interventions, and provide a foundation to assist in shared decision-making during procedure and patient selection.

## Data Availability

The National Inpatient Sample (NIS) offers the largest database of hospitalizations, including data on approximately 7-8 million discharges per year and representing a 20% random and stratified sample of hospital discharges in the United States, and all the data under NIS are publicly available.

https://hcup-us.ahrq.gov/nisoverview.jsp

## Acknowledgements

We would like to thank the Cardiovascular Department at Jackson Memorial Hospital and the University of Miami for their contributions in developing and completing this project.

